# Assessing the mental health literacy of young adults from rural and urban communities in Malawi

**DOI:** 10.1101/2024.12.03.24318388

**Authors:** Beatrice Cynthia Chitalah, Ishani Nanda, Joel Nyali, Gloria Chirwa, Sandra Jumbe

## Abstract

Mental health literacy is the ability to recognise mental disorders, have knowledge of professional help available, effective self-help strategies, prevention strategies, and skills to support others. Mental health literacy is linked to better help-seeking behaviours and the management of mental illness. The prevalence of mental illness in Malawi is increasing. Assessing mental health literacy in communities crucially helps identify knowledge gaps, informing the development of evidence-based interventions. This study assessed the mental health literacy levels of young adults (16-to-30 years old) in rural and urban communities in Malawi. A cross-sectional national survey was administered to 682 people across 13 districts in Malawi using a self-reporting Mental Health Literacy questionnaire (MHLq) that assessed knowledge of mental health problems, erroneous beliefs/stereotypes, first-aid skills, help-seeking behaviour, and self-help strategies. Most respondents were either unemployed (36%) or enrolled in school (43%). 73% had completed primary or secondary education, 48% knew someone with a mental illness but only 14.1% of this group could specify the illness. The mean mental health literacy score was 111.8 [Standard Deviation (SD) = 13.9]. Individuals with primary and secondary school qualifications had significantly lower scores in Factor 2 (erroneous beliefs/stereotypes) and Factor 3 (First-aid skills and assistance-seeking behaviour) of the MHLq than those with higher education. This research highlights persisting mental health misconceptions, limited knowledge about specific mental illnesses, and low help-seeking behaviour among young adult Malawians. Higher education is linked to a better understanding of mental health. Prioritising community education on causes, signs, treatments, and prognosis of mental illness is crucial for increased mental health literacy.

## Introduction

Mental health problems (MHPs) are becoming a significant issue among the youth population globally 1, with a 90% treatment gap in Southern African countries, including Malawi which does not align with the 10-30% prevalence of common mental disorders amongst young people alone 4,5. Approximately half of adult mental health disorder diagnoses start by age 14, impacting individuals’ functionality and productivity 2,3. Untreated MHPs can have debilitating effects on social life and increase vulnerability to chronic physical conditions 6. Research is being conducted to develop preventive approaches to reduce cases and promote mental wellbeing among young people.

Mental health literacy (MHL) is a significant determinant of mental health with the potential of improving health 7. It consists of five basic components: recognizing mental disorders, knowing available professional help, using effective self-help strategies, skills on supporting others, and mental disorders prevention knowledge 8. Sufficient MHL helps in understanding risk factors, and identifying common mental disorder symptoms, resulting in mental disorder prevention and reduced burden of disease 9. It also reduces stigma associated with mental illness, and informs better help-seeking intervention planning, thereby improving treatment-seeking attitudes and adherence among diagnosed cases.

Assessing MHL levels helps identify knowledge gaps and erroneous beliefs about mental health issues, which hinder early recognition and management 10. Recognizing these issues is crucial for developing interventions aimed at enhancing knowledge and challenging erroneous beliefs, ultimately promoting mental health 10.

### Study aim

Despite the increasing rates of mental health problems among youth in Malawi, there is limited documentation on programs targeting MHL 21. There is no study in Malawi assessing MHL among young adults in its community settings aside from our previous work on the Chichewa-translated MHL questionnaire 4. This study therefore aims to report outcomes from our national survey conducted in Malawi, which assessed the MHL levels of young adults in rural and urban communities.

## Methods

### Procedure

The study was approved by the National Committee on Research in the Social Sciences and Humanities (NCRSH) on 16 March 2021 before commencing with the current study (Ref. No. P12/20/539). A multistage sampling strategy was used, starting with a scoping meeting with our stakeholders Drug Fight Malawi (DFM) and the National Youth Council of Malawi (NYCOM) to identify districts across all three regions of Malawi to aid representative sampling of the general Malawi young adult population (16 to 30 years). Each district was allocated a quota based on variables such as geographical region , urban vs rural settings, tribal representation and socioeconomic status (see Figure 1). Surveying was conducted by fieldworkers from several youth led organisations and the research team (see Figure 1) between 25 March and 10 December 2021. Fieldworkers randomly approached young people in community hotspots like trading centres and sports grounds to complete a survey. In some districts (Nkhata Bay, Mzuzu, Phalombe), existing stakeholders working as youth workers were used as fieldworkers to better engage with youth and address language issues.

**Figure 1:**
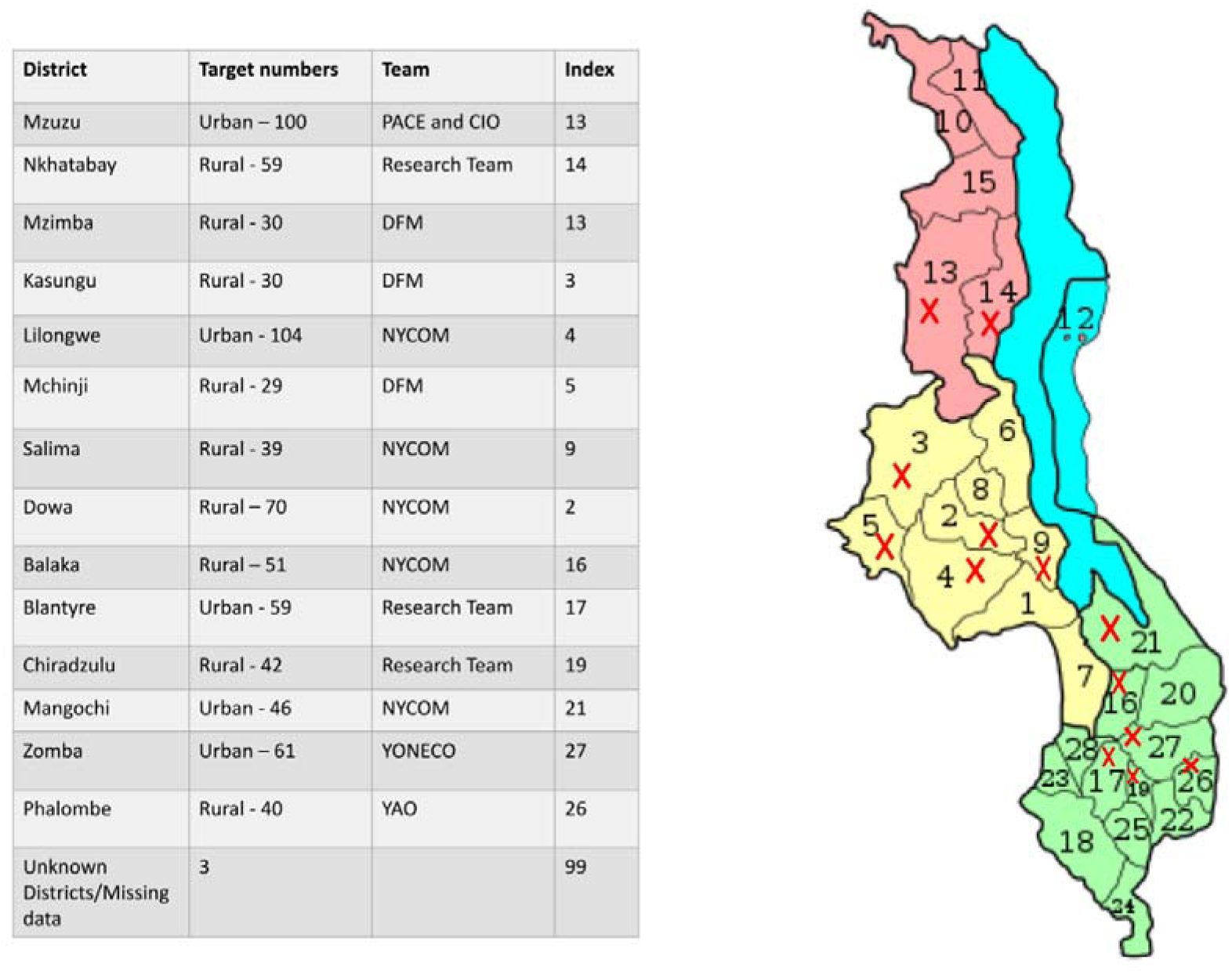
Overview of districts where Mental Health Literacy survey was conducted.

### Sample size

The survey was conducted among 16 to 30-year-old participants who were literate in English or Chichewa. Those unable to read and write independently were excluded.

Slovin’s formula23 was used to determine an appropriate sample size for this target population. Our sample size is based on the general population of young adults (16 – 30-year-olds) in Malawi, who make up a quarter of the country’s population, approximately 5 million people. We calculated the sample size using a 95% confidence interval, 5% margin of error and a standard deviation (SD) of 0.5. The SD value of 0.5 was because of uncertainty regarding how extreme the respondents’ answers would be. This gave a sample size of at least 384. We anticipated a low survey completion/ response rate of 50% because MHL survey study had not been done before and discussing mental health is culturally sensitive in Malawi. This gave a total sample size of 768.

### Data collection tool

A self-reporting MHL questionnaire (MHLq) for young adults which assesses four aspects of the MHL construct namely; knowledge about MHPs, erroneous beliefs/stereotypes, first-aid skills, help seeking behaviour and self-help strategies was used 10. Psychometric properties of the MHLq regarding validity and reliability have already been shown for this study population 10. We translated the questionnaire into Chichewa (local language) for use in rural communities, and to facilitate its use by mental health practitioners and researchers to screen mental health needs in diverse settings in Malawi. Preliminary reliability results for this Chichewa MHLq also suggest a reasonably good fit but more psychometric testing with larger samples is vital to further validate the questionnaire. 4.

Data collection followed ethical guidelines with all participants providing written informed consent prior to completing the survey. Participants completed part one of the questionnaire which consisted of questions related to the participant’s gender, age, marital status, nationality, residence, academic qualifications, profession/occupation, proximity to people with MHPs including the nature of the relationship (socio-demographics section). Then part two which contained 29 MHLq items based on the four aforementioned factors, organised in a 5-point Likert response scale (1=strongly disagree to 5 = strongly agree). For this sample, Total scores for the 29 items ranged between 15 and 145. The knowledge of MHPs factor scores ranged between 7 and 55 (mean score = 42.9; SD = 6.4). Erroneous beliefs/stereotypes factor scores (items 6, 10, 13, 15, 23, and 27 reverse scored)—ranged between 0 and 40 (mean score = 29.4; SD = 5.0). The first-aid skills and help seeking behaviour factor ranged between 4 and 30 (mean score = 23.0; SD = 4.8). Factor 4 scores regarding self-help strategies ranged between 4 and 20 (mean score = 16.4; SD = 2.7).

### Data analysis

Descriptive statistics were used to determine sample demographics, levels of missing data, and summarise participants’ MHL levels (Table 1). MHL scores by the four factors (sum of the values of the dimension items) and overall MHL scores (sum of the values of all the items) were computed. Higher values in all dimensions and overall score indicated higher levels of MHL 10. The relationship between MHL levels and socio-demographic variables (gender, geographical setting, language and proximity to someone with a mental health problem) was explored using t-tests for independent samples. A one-way analysis of variance (ANOVA) was used to examine the relationship between education levels (primary, secondary, and tertiary) and MHL scores. A Pearson’s correlation test was used to assess the relationship between overall MHL score (including four MHLq factor scores) and age.

**Table 1:** Survey respondents’ characteristics.

| **Table 1: Survey respondents' characteristics** |  |  |  |
| --- | --- | --- | --- |
| **Sociodemographic variables**<br>*(missing value)* |  | **n** | **Valid %** |
| **Gender**<br>*(56)* | Female | 336 | 54 |
|  | Male | 290 | 46 |
| **Mean age (SD)** |  | 21 (3.48) |  |
| **Occupation**<br>*(19)* | Employed full time | 66 | 9.7 |
|  | Employed part-time | 63 | 9.2 |
|  | Unemployed | 243 | 35.6 |
|  | Student | 291 | 42.6 |
| **Marital status**<br>*(28)* | Single | 519 | 76.1 |
|  | Married | 117 | 17.2 |
|  | Divorced | 10 | 1.5 |
|  | Widowed | 8 | 1.2 |
| **Education**<br>*(33)* | Completed primary | 244 | 35.8 |
|  | Completed secondary | 253 | 37.1 |
|  | Higher | 152 | 22.3 |
| **Geographical setting**<br>(0) | Urban | 282 | 41.3 |
|  | Rural | 400 | 58.7 |
| **Knowing someone with a mental health problem**<br>(8) | Yes | 324 | 47.5 |
|  | No | 287 | 42.1 |
|  | Not sure | 60 | 8.8 |
| **Proximity to person with a mental health problem**<br>(24) | Friend | 105 | 30.1 |
|  | Myself | 9 | 2.6 |
|  | Relative | 59 | 16.9 |
|  | Someone else | 152 | 22.3 |
| **Language survey completed in** | Chichewa | 525 | 77.0 |
|  | English | 157 | 23.0 |

Lastly, responses to the open-ended question ‘What type of mental health problem?’ were qualitatively coded for those who ticked ‘Yes’ to the baseline question “Do you know anyone who has or had a mental health problem?” This was done to gauge the types of MHPs people within participating communities we re aware of. Missing data was computed as ‘99’ in the whole dataset. In all hypothesis tests, a level of significance of 5% was considered. All analyses were performed using the statistical analysis program IBM Statistical Package for Social Sciences (SPSS) version 24.0.

## Results

A total of 763 participants completed the questionnaire from 13 districts in Malawi (see figure 1) . Table 1 describes demographic characteristics of the 682 survey participants with complete data, after excluding those with missing ages, those outside the age range of 16 to 30 years and those who did not respond to any MHLq items (n=7). Of the young adults, 54% were female, and 76% were single. Regarding occupation of respondents, most participants were students (43%) but another large chunk were unemployed (36%). Most participants’ (72.9%) highest educational attainment was either primary (35.8%) or secondary (37.1%) education. 325 respondents (47.6%) reported knowing someone who had a mental health problem. Regarding proximity to said person with a mental health problem, 152 of the 325 respondents said it was someone not close to them. The others stated that they had a relative (n=59 or friend (n=105) with a mental health problem. Only nine respondents stated that they themselves had a mental health problem. 525 out of the 682 respondents (77%) completed the survey in Chichewa language.

### Mental Health Literacy scores and sociodemographic variables

The mean MHL total score for this sample was 111.8 (SD = 13.9). Independent t-test results showed significant differences in MHLq scores between groups for the socio-demographic variable, language (see Table 2). Those who responded in English had higher total MHLq scores (116.0, ± 12.9) than Chichewa respondents (110.5, ± 14.0), t(680) = -4.382, p <0.001, along with all MHLq factors, except for Factor 1 (Knowledge of MHPs ), where no significant difference was found. However, homogeneity of variances assumption was not met for factors 3 (First-aid skills and help seeking behaviour) and 4 (Self-help strategies), indicating substantial differences in variances among groups therefore Welch test outcomes are reported instead.

**Table 2.** Differences in mental health literacy (MHLq global score and dimensions) based on language used to complete survey.

|  | Chichewa<br>(n= 525) | English<br>(n=157) | t value<br>(df=680) |
| --- | --- | --- | --- |

|  | Mean (SD) | Mean (SD) |  |
| --- | --- | --- | --- |
| MHL (total score) | 110.5 (13.9) | 116.0 (12.9) | -4.382*** |
| Knowledge of MHPs<sup>a</sup> | 42.8 (6.5) | 43.4 (5.9) | -1.014 |
| Erroneous beliefs/stereotypes | 28.8 (4.6) | 31.6 (5.3) | -6.447*** |
| First-aid skills and help seeking behaviour<sup>a</sup> | 22.7 (5.0) | 24.1 (4.0) | -3.841*** |
| Self-help strategies<sup>a</sup> | 16.3 (2.8) | 16.9 (2.3) | -2.754** |
| * p < 0.05; ** p < 0.01; *** p < 0.001 |  |  |  |
| <sup>a</sup>Welch test reported as homogeneity of variances assumption was not met for this variable |  |  |  |

Differences in MHL scores were also noted between participants from rural and urban settings. Rural respondents had significantly higher mean scores than urban participants on the MHLq total score, knowledge of MHPs , erroneous beliefs/stereotypes and self-help strategies (Table 3). No significant difference in mean scores was noted for Factor 3 (First-aid skills and help seeking) between groups. Interestingly, this was the only factor where rural participants had a mean score that was very slightly lower than urban participants. No significant differences in MHLq scores were noted for the other variables (gender and knowledge of someone with MHPs ).

**Table 3.** Differences in mental health literacy (MHLq global score and dimensions) between rural vs urban settings.

|  | Rural<br>(n= 400)<br>Mean (SD) | Urban<br>(n=282)<br>Mean (SD) | t value<br>(df=680) |
| --- | --- | --- | --- |
| MHL (total score)<sup>a</sup> | 113.0 (12.7) | 110.2 (15.3) | 2.494* |
| Knowledge of MHPs<sup>a</sup> | 43.5 (6.0) | 42.2 (6.8) | 2.671** |
| Erroneous beliefs/stereotypes<sup>a</sup> | 29.8 (4.5) | 29.0 (5.5) | 2.041* |
| First aid skills and help seeking behaviour | 23.0 (4.9) | 23.1 (4.6) | -.230 |
| Self-help strategies<sup>a</sup> | 16.7 (2.5) | 16.0 (3.0) | 3.213*** |
| * p < 0.05; ** p < 0.01; *** p < 0.001 |  |  |  |
| <sup>a</sup>Welch test reported as homogeneity of variances assumption was not met for this variable |  |  |  |

The relationship between education levels (primary, secondary and tertiary) and MHL scores was examined using one-way analysis of variance (ANOVA). However, these failed to meet Levene’s test of homogeneity assumption, meaning there were substantial differences in variances among groups. Therefore, the Welch test (Table 4) was conducted along with the Games-Howell post hoc test (Table 5). There was no statistically significant difference noted between education groups for the total MHL score (p = 0.192). Statistically significant differences between education groups were noted for factor 2 ‘Erroneous beliefs & stereotypes’ (F(2, 646) = 3.911, p = 0.021) and factor 3 ‘First aid skills and help seeking behaviour’ (F(2, 646) = 7.179, p < 0.001).

**Table 4:**
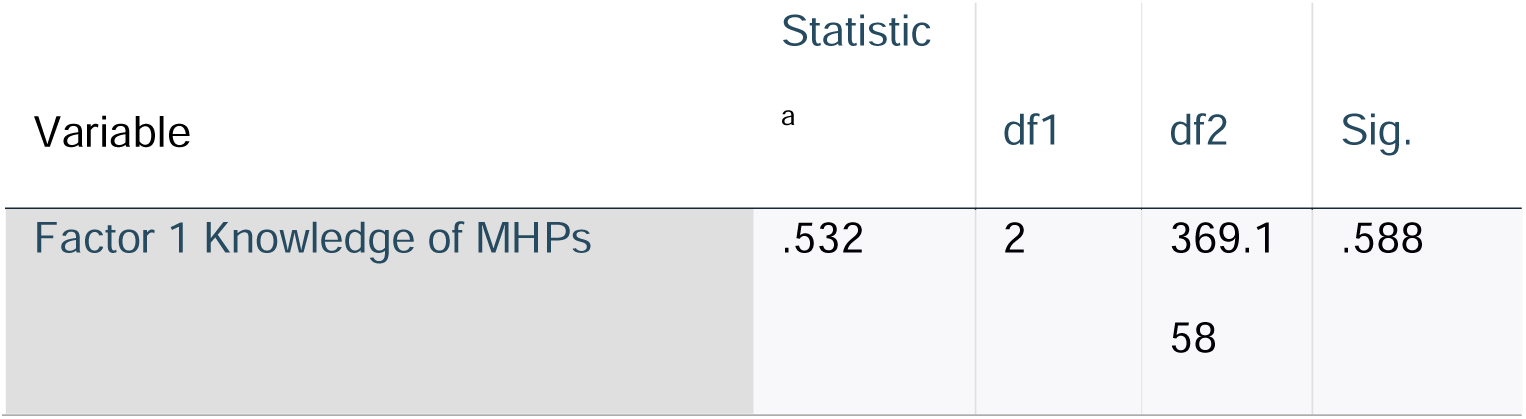
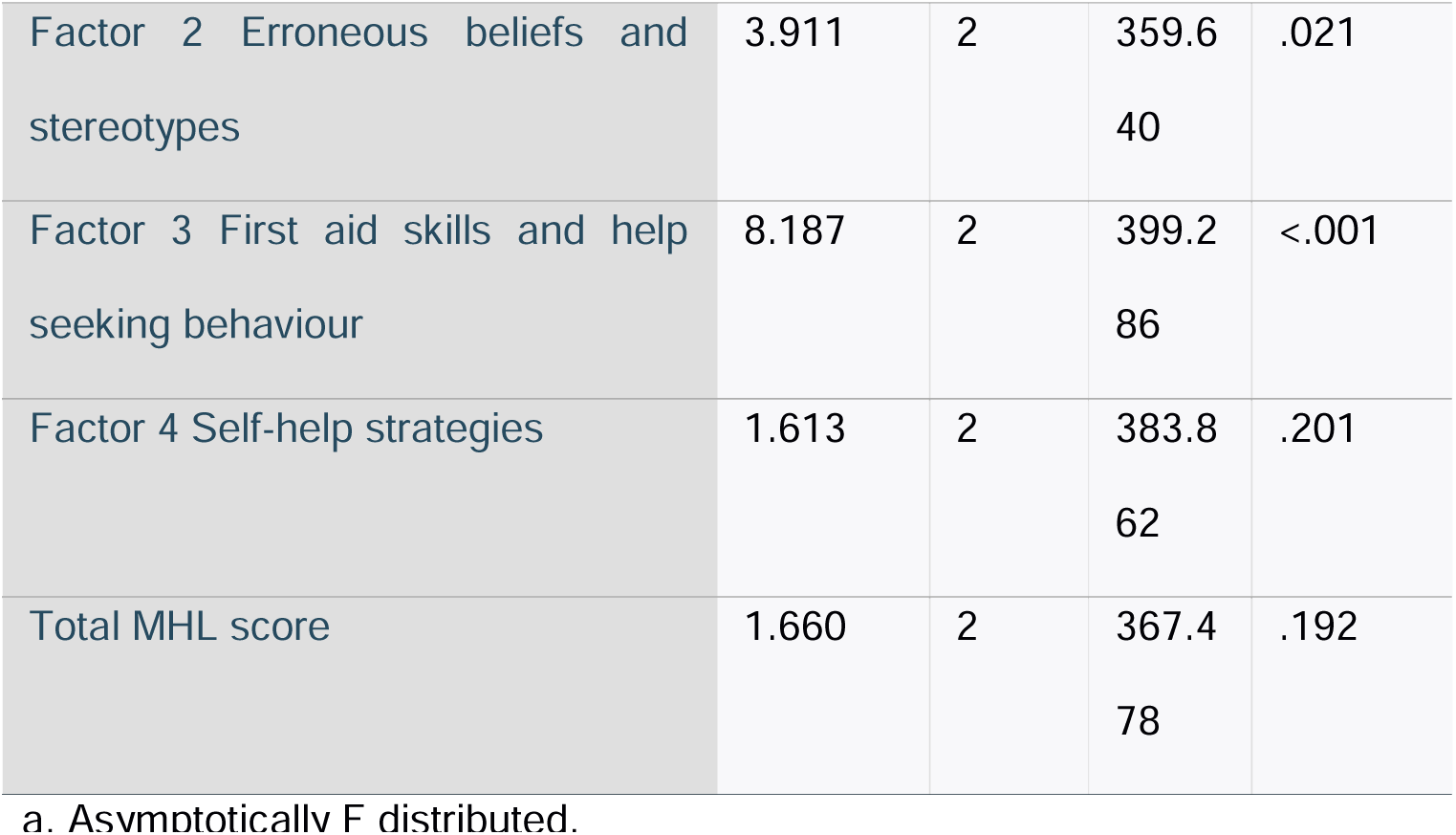
Welch Robust Tests of Equality of Means.

**Table 5:**
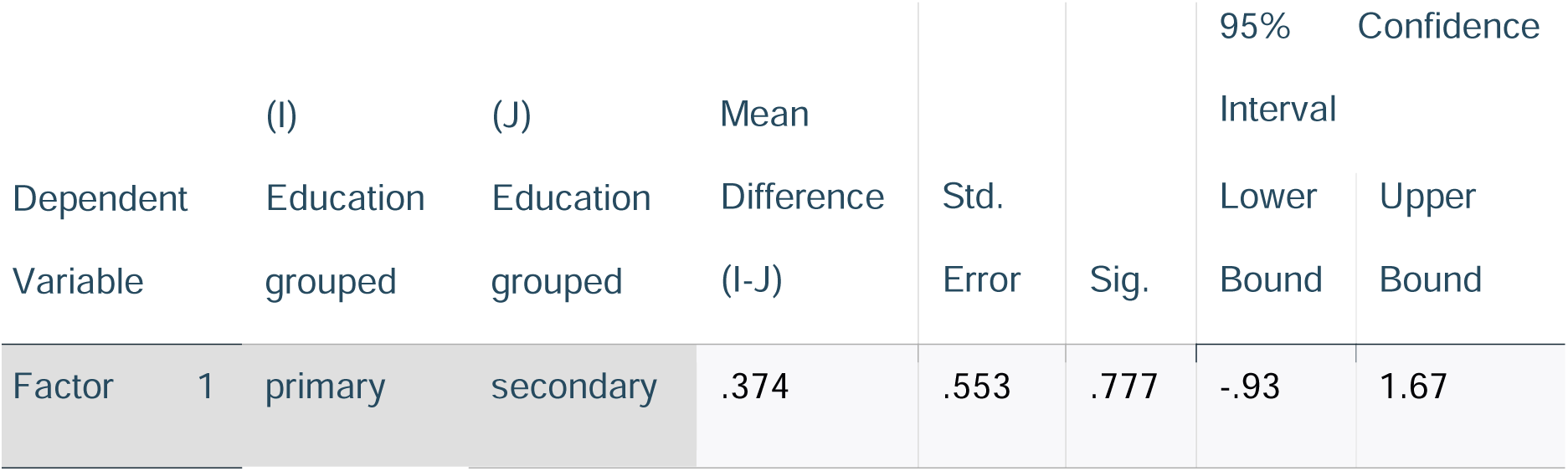
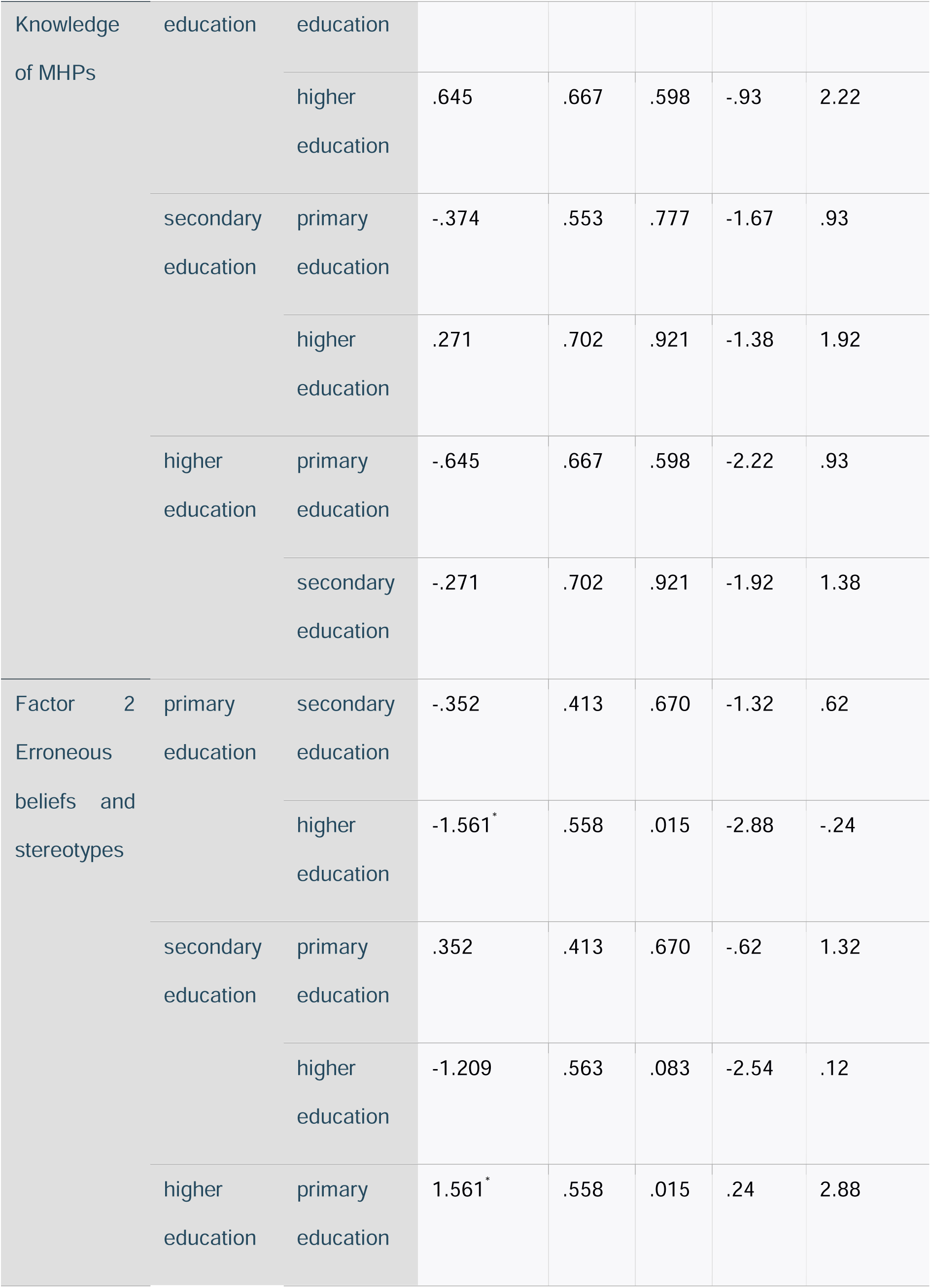
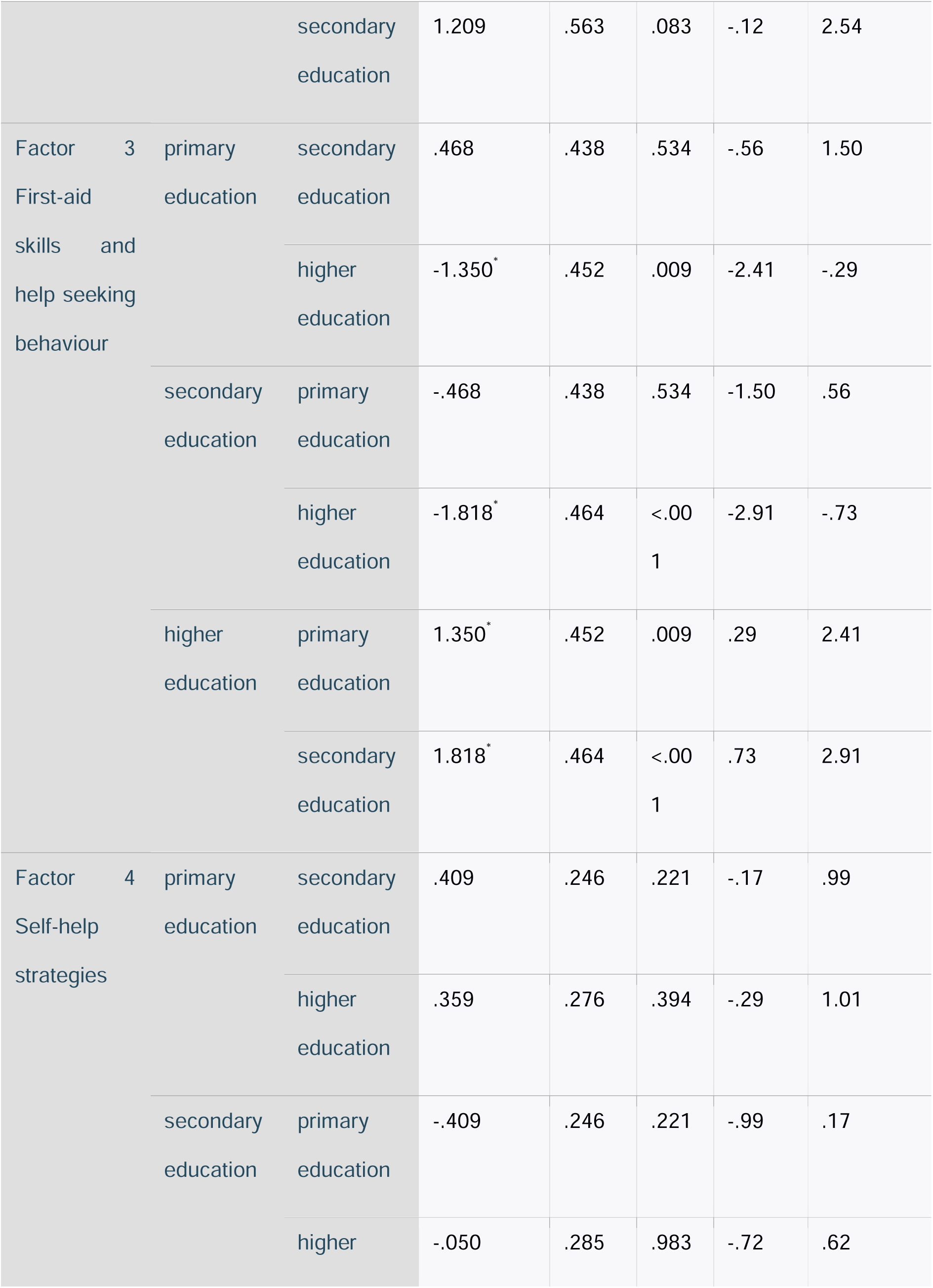
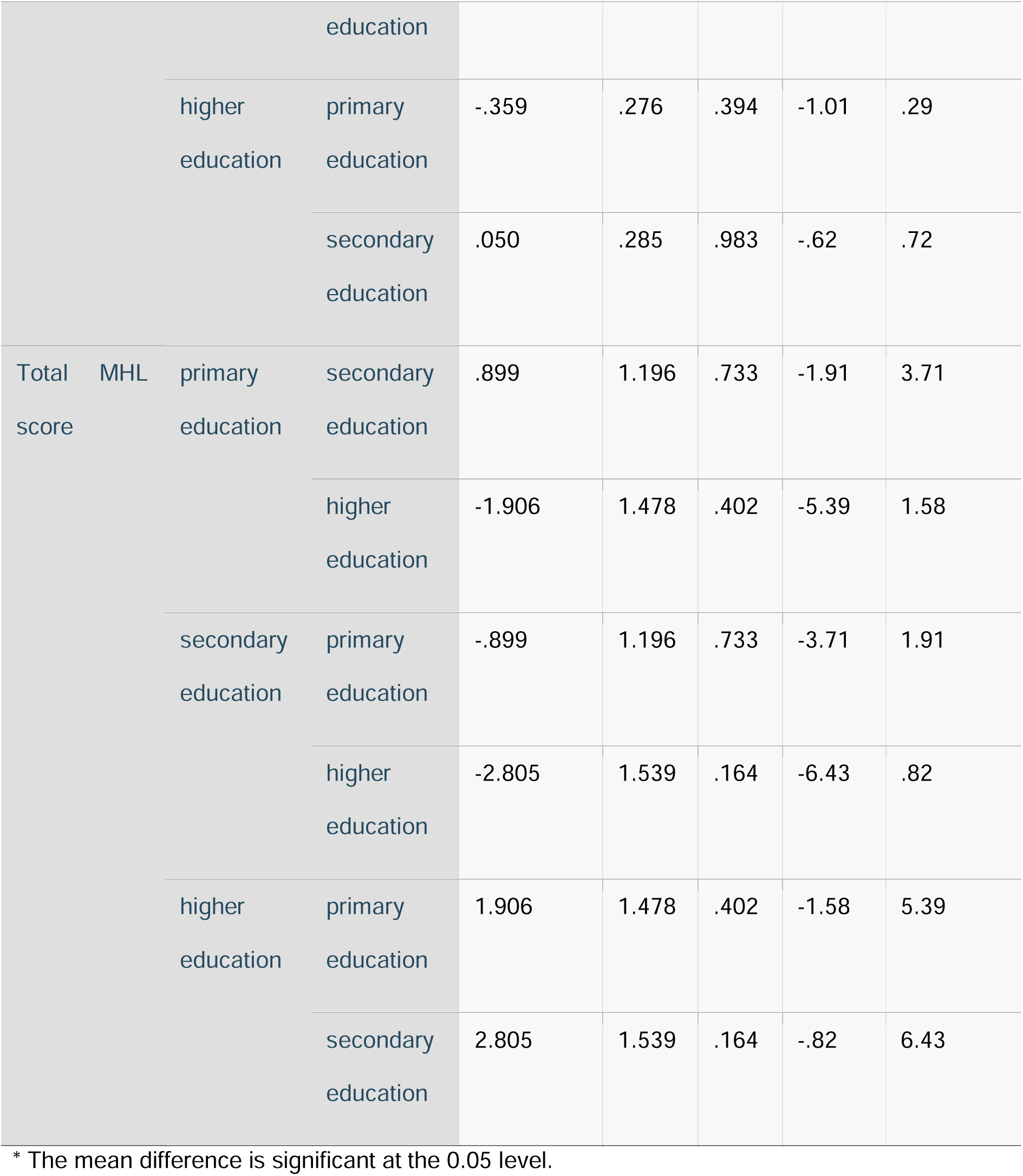
Multiple Comparisons using Games-Howell post hoc test.

Regarding factor 2, the Games-Howell post hoc test revealed that MHL factor 2 scores were statistically significantly lower for those who had completed primary school (29.0 ± 4.5) compared to those who had completed higher education (30.6 ± 5.9). There was no statistically significant difference between the primary and secondary groups (p = .670) or the secondary and higher education groups (*p* = 0.083). For factor 3, there was similarly no statistically significant difference between the primary and secondary groups (p = 0.534). However, MHL factor 3 scores were statistically significantly lower for those who had completed primary school (22.9 ± 4.7, *p* = 0.009) and secondary school (22.5 ± 5.1, *p* < 0.001) compared to those in higher education (24.3 ± 4.2).

Taken together, these results suggest that education has an effect on MHL scores. Specifically, our results suggest that those who go on to study at university or college or have higher education qualifications have a better understanding of mental health than those with primary or secondary school qualifications. It should be noted that there doesn’t seem to be much difference in MHL between those who have completed primary and secondary education.

A Pearson’s correlation test was used to assess the linear relationship between MHL score (including four MHLq factor scores) and age (Table 6). There was a significant but very weak negative relationship between the MHL total score and age variables, r(680) = -.088, p = 0.022. In terms of the MHLq factor scores, there was also a significant negative relationship between the factor 2 ‘Erroneous beliefs and stereotypes’ scores and age, r(680) = -.083, p = 0.029. No significant relationship was noted with the other 3 MHLq factors. Overall results suggest that younger adults may have higher MHL than older adults.

**Table 6:** Correlations between MHL scores and age.

|  |  | Age |
| --- | --- | --- |
| Factor 1<br>Knowledge of MHPs | Pearson | -.062 |
|  | Correlation |  |
|  | Sig. (2-tailed) | .105 |
|  | N | 682 |
| Factor 2<br>Erroneous beliefs and stereotypes | Pearson | -.083<sup>*</sup> |
|  | Correlation |  |
|  | Sig. (2-tailed) | .029 |
|  | N | 682 |
| Factor 3<br>First-aid skills and help seeking<br>behaviour | Pearson | -.045 |
|  | Correlation |  |
|  | Sig. (2-tailed) | .237 |
|  | N | 682 |
| Factor 4 | Pearson | -.071 |
| Self-help strategies | Correlation |  |
|  | Sig. (2-tailed) | .062 |
|  | N | 682 |
| Total MHL score | Pearson | -.088* |
|  | Correlation |  |
|  | Sig. (2-tailed) | .022 |
|  | N | 682 |
| * p < 0.05; ** p < 0.01; *** p < 0.001 |  |  |

### Types of mental health problems

Out of the 325 individuals who disclosed knowing someone with an MHP , only 14.1% were able to identify what the specific mental health problem of the person they knew was. In particular, they reported knowing someone with conditions such as PTSD, (post-traumatic stress disorder), anxiety, stress, schizophrenia and bipolar (Table 7). 34.2% wrote terms like mental disorder, brain problem (‘vuto la ubongo’ and ‘mutu sugwira ntchito’ are the Chichewa equivalents) or madness (misala, kupenga and kuzungulira mutu are the Chichewa equivalents), without being exact on the type of mental disorder. Seven individuals mentioned epilepsy, a neurological condition. The rest of the participants (31%) were unable to identify specific MHPs or disorders, instead, what they mentioned were more of potential signs and symptoms (e.g. wandering about aimlessly, talking to self) or antisocial behaviours (e.g. stealing, fighting).

**Table 7.**
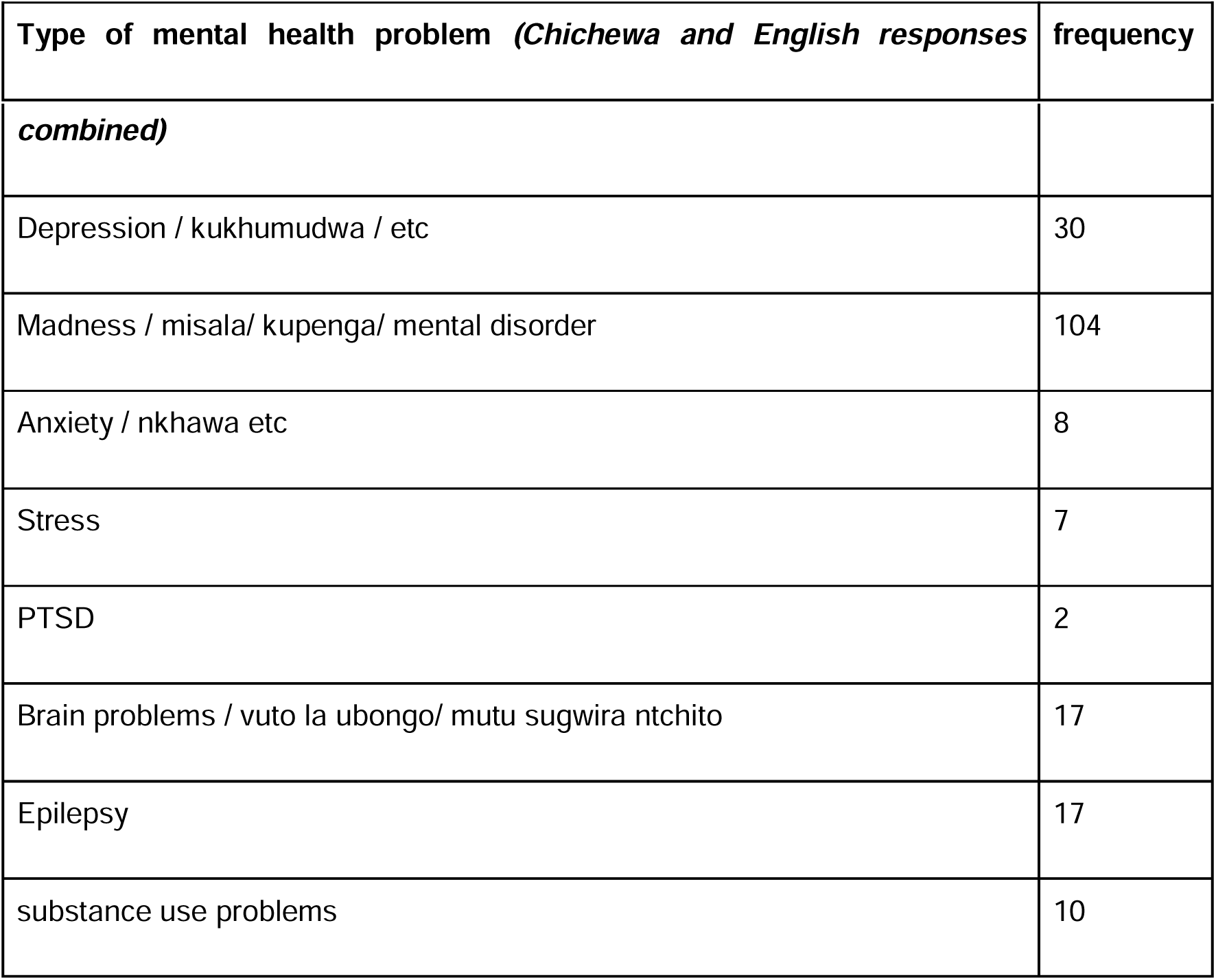
Types of mental health problems reported by respondents.

## Discussion

A majority of people have a higher chance of developing mental health disorders due to the global increase in mental health illnesses during a lifetime8. This study aimed to assess MHL levels among young adults in Malawi, focusing on rural and urban settings. The survey revealed that 47.6% of respondents knew someone with mental health issues, but only 14% could identify the specific disorder. This lack of identification can lead to delayed treatment and intervention, reduced quality of life, and increased burden on families and caregivers14. The study also found significant differences in mental health literacy levels between education groups for “Erroneous ideas and stereotypes” (F(2, 646) = 3.911, p = 0.021) and “First-aid skills and assistance-seeking behaviour” (F(2, 646) = 7.179, p 0.001). The higher one’s education level, the higher one’s MHL level. There was however no significant difference in MHL scores between primary and secondary education participants. Higher education levels resulted in a lower likelihood of supporting negative ideas or stigmatising notions15.

A study in Malawi by Chilale et al.17 found that understanding and knowledge of mental health illnesses with biopsychosocial inclination can increase help-seeking behaviour. The prevailing socio-cultural explanation of witchcraft and spirit possession influences people’s behaviour when seeking treatment. Participants acknowledged the benefits of hospitalisation but also acknowledged difficulties, including biopsychosocial aspects. The study suggests reinforcing a biopsychosocial intervention paradigm for mental disorder treatment. There is also a need for anti-stigma teaching initiatives in Sub-Saharan Africa, where societal separation and stigmatisation of mental illness are prevalent. Therefore, incorporating such initiatives into mental health policies is crucial16. Our study primarily consisted of a sample of young adults, just over half were women, 76% were single and a third unemployed. Most participants completed primary or secondary education. Women make up 52% of the total Malawi population13 however unemployment in our sample was higher than the national estimate of 9%20.

This study has several strengths. This study is the first in Malawi to assess mental health literacy among young adults in community settings. With a large sample size of 682 participants, the findings are generalis able to the young adult population aged 16-30 in Malawi. The sample representativeness was well considered due to a sampling strategy accounting for key variables including participants from both urban and rural areas across the country. The open-ended questions in the survey provided crucial depth to the data. The cross-sectional survey approach was cost effective and efficient. Having both English and Chichewa versions of the MHLq made the survey accessible to a wider range of individuals in both rural and urban communities.

Study limitations include its focus on young adults (16-30 years old) which limits its applicability to different ages in the community. We had to exclude 81 respondents from our analyses due to missing data. Lacking r esources and time meant we could not recoup this lost data. In future, we will need to have a strategic protocol in place with fieldworkers to minimise missing data.

To conclude, the study aimed to assess mental health knowledge and attitudes among young adults, highlighting gaps in education. The survey emphasised the need for educational interventions targeting MHL at a younger age, particularly in primary and secondary schools, to reduce stereotyping and negative/stigmatising beliefs towards individuals with MHP, enabling them to seek professional help more effectively. Community education on mental health should focus on underlying factors, symptoms, treatment, and prognosis. Schools should provide mental health education with treatment and aid options to reduce stereotypes 8,11. Research shows that providing an MHL education curriculum improves student knowledge of mental illness, reduces stigmatising attitudes, and makes it easier to seek assistance and identify problems early. This approach increases the success rate of treatments19, as individuals who can identify their own or family members’ mental health issues may receive prompt help and treatment14,8.

The survey data can be used by mental health practitioners and academics to promote MHL in rural and urban settings, especially among disadvantaged groups. Adequate MHL is crucial for early detection and treatment of mental health issues and promoting mental wellness. However, there is limited research on MHL in developing countries like Malawi12. More longitudinal studies are needed to understand its changes over time and to translate data collection tools into native languages. Culturally relevant MHL programs should be delivered across communities, considering individual literacy levels.

## Data Availability

All data produced in the present study are available upon reasonable request to the authors

## Acknowledgements

The authors are incredibly grateful to all the participants who took part in this work. We also thank our project partners and stakeholders; National Youth Council of Malawi, Drug Fight Malawi, Youth Net Counselling, Pace for change, Phalombe Youth Arms Organisation, Chimwemwe Banda and Gibson Kaunda for their invaluable help and support. This research was supported by the African Academy of Sciences. The funder was not involved in study design, data collection, data analysis, or manuscript preparation.

## References

1. Jurewicz I. Mental health in young adults and adolescents - supporting general physicians to provide holistic care. Clin Med J. 2015;15(2):151. doi:10.7861/clinmedicine.15-2-151

2. Solmi M, Radua J, Olivola M, Croce E, Soardo L, Salazar de Pablo G, et al. Age at onset of mental disorders worldwide: large-scale meta-analysis of 192 epidemiological studies. Mol. Psychiatry. 2021;27(1). https://www.nature.com/articles/s41380-021-01161-7

3. Otto, C., Reiss, F., Voss, C., Wüstner A., Meyrose A., Hölling H., et al. Mental health and well-being from childhood to adulthood: design, methods and results of the 11- year follow-up of the BELLA study. Eur Child Adolesc Psychiatry. 2021;30:1559–1577. doi:10.1007/s00787-020-01630-4

4. Jumbe S, Nyali J, Newby C. Translation of the mental health literacy questionnaire for young adults into Chichewa for use in Malawi: preliminary validation and reliability results. Int J Ment Health Syst. 2023 Jun 8;17(1). doi:10.1186/s13033-023-00586-7

5. Chilale HK, Silungwe ND, Gondwe S, Masulani-Mwale C. Clients and carers perception of mental illness and factors that influence help-seeking: Where they go first and why. International Journal of Social Psychiatry. 2017;12;63(5):418–25. doi:10.1177/0020764017709848

6. Kaushik A, Kostaki E, Kyriakopoulos M. The stigma of mental illness in children and adolescents: A systematic review. Psychiatry Res. 2016;243:469–94. https://pubmed.ncbi.nlm.nih.gov/27517643/

7. Wei Y, McGrath PJ, Hayden J, Kutcher S. Mental health literacy measures evaluating knowledge, attitudes and help-seeking: a scoping review. BMC Psychiatry, 2015; 15, 291. doi:10.1186/s12888-015-0681-9

8. Jorm AF. Mental health literacy: empowering the community to take action for better mental health. Am. Psychol. 2012;67(3):231–43. doi:10.1037/a0025957

9. Sequeira C, Sampaio F, Pinho LG, Araújo O, Lluch Canut T, Sousa L. Editorial: mental health literacy: how to obtain and maintain positive mental health. Front Psychol. 2022 Oct 12;13. doi:10.3389/fpsyg.2022.1036983

10. Dias P, Campos L, Almeida H, Palha F. Mental health literacy in young adults: adaptation and psychometric properties of the mental health literacy questionnaire. Int J Environ Res Public Health. 2018 Jun 23;15(7):1318. doi:10.3390/ijerph15071318

11. Kutcher S, Gliberds H, Morgan C, Greene R, Hamwaka P, Perkins K. Improving Malawian teachers’ mental health knowledge and attitudes: an integrated school mental health literacy approach. Camb Prism. 2015 Feb 16; 2:e1. doi:10.1017/gmh.2014.8

12. Nobre J, Oliveira AP, Monteiro F, Sequeira C, Ferré-Grau C. Promotion of mental health literacy in adolescents: a scoping review. Int J Environ Res Public Health. 2021 Sep 9;18(18):9500. doi:10.3390/ijerph18189500

13. Statista. Malawi - total population 2018-2028 | Statista. Available from: https://www.statista.com/statistics/520538/total-population-of-malawi/.

14. Dutta M, Spoorthy MS, Patel S, Agarwala N. Factors responsible for delay in treatment seeking in patients with psychosis A qualitative study. Indian J Psychiatry. 2019;61(1):53–9. doi:10.4103/psychiatry.IndianJPsychiatry_234_17

15. Hartini N, Fardana NA, Ariana AD, Wardana ND. Stigma toward people with mental health problems in Indonesia. Psychol. res. behav. 2018;11(11):535–41. 10.2147/PRBM.S175251

16. Anosike C, Aluh D, Onome O. Social Distance Towards Mental Illness Among Undergraduate Pharmacy Students in a Nigerian University. East Asian Arch Psychiatry 2020;30:57–62. doi:10.12809/eaap1924

17. Chilale HK, Silungwe ND, Gondwe S, Masulani-Mwale C. Clients and carers perception of mental illness and factors that influence help-seeking: where they go first and why. Int J Soc Psychiatry. 2017 Jun 12; 63(5):418–25. doi:10.1177/0020764017709848

18. Jumbe S, Nyali J, Simbeye M, Zakeyu N, Motshewa G, Pulapa SR. ‘We do not talk about it’: Engaging youth in Malawi to inform adaptation of a mental health literacy intervention. Plos One. 2022 Mar 29;17(3):e0265530. doi:10.1371/journal.pone.0265530

19. Beukema L, Tullius JM, Korevaar L, Hofstra J, Reijneveld SA, de Winter AF. Promoting mental health help-seeking behaviours by mental health literacy interventions in secondary education? Needs and perspectives of adolescents and educational professionals. Int J Environ Res Public Health. 2022 Sep 20;19(19):11889. doi:10.3390/ijerph191911889

20. Chinsinga B, Chasukwa M. Agricultural policy, employment opportunities and social mobility in rural malawi. Agrar South. 2018 Mar 29;7(1):28–50. doi: 10.1177/2277976018758077

21. Kauye F, Udedi M, Mafuta C. Pathway to care for psychiatric patients in a developing country: Malawi. Int J Soc Psychiatry. 2014 Jun 4. doi:10.1177/0020764014537235

22. Malone HE, Nicholl H, Coyne I. Fundamentals of estimating sample size. Nurse Res. 2016 May;23(5):21–5. doi: 10.7748/nr.23.5.21.s5.

